# Genomic surveillance of *Clostridioides difficile* transmission and virulence in a healthcare setting

**DOI:** 10.1101/2023.09.26.23295023

**Authors:** Erin P. Newcomer, Skye R. S. Fishbein, Kailun Zhang, Tiffany Hink, Kimberly A. Reske, Candice Cass, Zainab H. Iqbal, Emily L. Struttmann, Erik R. Dubberke, Gautam Dantas

## Abstract

*Clostridioides difficile* infection (CDI) is a major cause of healthcare-associated diarrhea, despite the widespread implementation of contact precautions for patients with CDI. Here, we investigate strain contamination in a hospital setting and genomic determinants of disease outcomes. Across two wards over six months, we selectively cultured *C. difficile* from patients (n=384) and their environments. Whole-genome sequencing (WGS) of 146 isolates revealed that most *C. difficile* isolates were from clade 1 (131/146, 89.7%), while only one isolate of the hypervirulent ST1 was recovered. Of culture-positive admissions (n=79), 19 (24%) of patients were colonized with toxigenic *C. difficile* on admission to the hospital. We defined 25 strain networks at ≤ 2 core gene SNPs; 2 of these networks contain strains from different patients. Strain networks were temporally linked (p<0.0001). To understand genomic correlates of disease, we conducted WGS on an additional cohort of *C. difficile* (n=102 isolates) from the same hospital and confirmed that clade 1 isolates are responsible for most CDI cases. We found that while toxigenic *C. difficile* isolates are associated with the presence of *cdtR*, nontoxigenic isolates have an increased abundance of prophages. Our pangenomic analysis of clade 1 isolates suggests that while toxin genes (*tcdABER and cdtR*) were associated with CDI symptoms, they are dispensable for patient colonization. These data indicate toxigenic and nontoxigenic *C. difficile* contamination persists in a hospital setting and highlight further investigation into how accessory genomic repertoires contribute to *C. difficile* colonization and disease.

## Background

*Clostridioides difficile* infection (CDI) is one of the most common healthcare-associated infections (HAIs) in the US and is the leading cause of healthcare-associated infectious diarrhea^1,2^. Since the early 2000s, *C. difficile* research has focused largely on hypervirulent strains, such as PCR ribotype 027^1,3–6^, which were responsible for hospital-associated CDI outbreaks. Strains of ribotype 027 were responsible for 51% and 84% of CDI cases in the US and Canada in 2005, respectively^1,4,5^. Since then, other circulating strains have emerged as the prevalent strains causative of CDI, such as 078 and 014/020^7–9^. One report indicated that the prevalence of PCR ribotype 027 decreased from 26.2% in 2012 to 16.9% in 2016^9^. As the landscape of *C. difficile* epidemiology continues to evolve, we must update our understanding of how various strains of this pathogen evolve, spread, and cause disease.

In addition to the changing prevalence of CDI-causing *C. difficile* strains, their transmission dynamics also appear to be evolving. In the late 1980s, it became clear that patients with active CDI shed spores onto their surroundings, leading to *C. difficile* transmission and future CDI events in the healthcare setting^1^. Because of this, patients with active CDI are placed on contact precautions to prevent transmission to susceptible patients, which has been successful in reducing rates of CDI^2,10^. Nevertheless, while epidemiological estimates indicate that 20-42% of infections may be connected to a previous infection, multiple genomic studies fail to associate a CDI case to a previous case^11–13^. This suggests other potential sources of disease development in the hospital environment. Indeed, while asymptomatic carriers of *C. difficile* have not been a significant focus of infection prevention efforts, studies have shown these carriers do shed toxigenic *C. difficile* spores to their surroundings that could cause disease^14^. Though carriers have not been consistently identified as major transmitters of *C. difficile* that causes CDI, recent work has suggested that patients carrying *C. difficile* asymptomatically may be at elevated risk for development of CDI^15^. Correspondingly, it is critical to both confirm this finding in another setting, and understand the genomic factors that may influence the transition from carrier to CDI manifestation in hospitalized patient populations. Correspondingly, it is critical to understand if *C. difficile* carriers are major contributors to new *C. difficile* acquisition or CDI manifestation in hospitalized patient populations.

*C. difficile* strains are categorized into five major clades and three additional cryptic clades. These clades encompass immense pangenomic diversity with many mobilizable chromosomal elements^16,17^, including numerous temperate phages that have potential influences over *C. difficile* toxin expression, sporulation, and metabolism^18^. Two major toxin loci, not required for viability, encode large multi-unit toxins that independently augment the virulence of *C. difficile*. Epithelial destruction and CDI have largely been attributed to the presence of pathogenicity locus (PaLoc) encoding toxins TcdA and TcdB. In addition, an accessory set of toxins (CdtA and CdtB) encoded at the binary toxin locus, may worsen disease symptoms^19^. Yet, many nontoxigenic strains of *C. difficile* have been documented and are adept colonizers of the GI tract, even without the PaLoc^20^. As there has been continued debate about strain-specific virulence attributes^21–23^, it is important to investigate the extent of strain-level pangenomic diversity and consequences of such diversity on host disease^24,25^.

The purpose of this study was to evaluate the role of *C. difficile* strain diversity in colonization outcomes and hospital epidemiology. By sampling patients (n=384) and their environments for six months in two leukemia and hematopoietic stem cell (HCT) transplant wards at Barnes-Jewish Hospital in St. Louis, USA, we used isolate genomics to identify environmental contamination of both toxigenic (TCD) and nontoxigenic (NTCD) *C. difficile* by carriers and CDI patients, and corresponding transmission between both patient groups. Integration of isolate genomic data and CDI information from this prospective study with isolate genomic data from a complementary retrospective study of asymptomatic vs symptomatic *C. difficile* colonization in the same hospital^26,27^ indicated that the clade 1 lineage, containing both toxigenic strains and nontoxigenic strains, dominates circulating populations of *C. difficile* in this hospital. Further, this lineage revealed novel clade-specifc genetic factors that are associated with CDI symptoms in patients.

## Methods

### Study Design

This prospective observational study took place in the leukemia and hematopoietic stem cell transplant (HCT) wards at Barnes-Jewish Hospital (BJH) in St. Louis, Missouri, United States. Each ward consisted of two wings with 16 beds; on the acute leukemia ward we enrolled from both wings (32 beds) and on the HCT ward we enrolled on one wing (16 beds). The wards were sampled for 6 months from January 2019-July 2019 (acute leukemia) and 4 months from March 2019-July 2019 (HCT). These units are located 2 floors apart in the same building. Colonized on admission was defined as: 1. Having a *C. difficile* culture positive specimen collected before calendar day 3 of admission, or 2. Having a history of an earlier *C. difficile* culture positive specimen of the same strain collected during a previous hospitalization. Acquisition was considered indeterminate when the earliest culture positive specimen was collected on calendar day 4 or later during admission.For EIA positive admissions, new acquisitions were defined as having toxigenic culture negative specimen that preceded the EIA positive clinical stool collection during the same admission. EIA positive admission were classified as indeterminate if the patient did not have any stool or rectal swab culture results available prior to the collection of the EIA positive stool sample. EIA positive admissions were defined as colonized on admission or pre-existing colonization if the patient had a toxigenic culture positive specimen that preceded the EIA positive clinical stool during the same admission.

### Sample collection, selective culture, and isolate identification

Patients and their environments were sampled upon admission to a study ward and then weekly until discharge. Per hospital standards, bleach is used for daily and terminal discharge cleaning. From each patient, a stool specimen and/or rectal swab was collected as available. Remnant fecal samples from the BJH microbiology laboratory that were obtained during routine clinical care for *C. difficile* testing were also collected. Stool samples and rectal swabs collected on enrollment were refrigerated for up to 3 hours before processing. Specimens from all other timepoints were stored in at -80°C in tryptic soy broth (TSB)/glycerol before processing. Environmental samples were collected from bedrails, keyboards, and sink surfaces using 3 E-swabs (Copan). If a surface was unable to be sampled, a swab was taken from the IV pump or nurse call button as an alternative. Swab eluate were stored at -80°C until processing.

Broth enrichment culture for *C. difficile* in Cycloserine Cefoxitin Mannitol Broth with Taurocholate and Lysozyme (CCMB-TAL) (Anaerobe Systems, Morgan Hill, CA) was performed on all admission specimens and checked for growth at 24 hours, 48 hours, and 7 days after inoculation. If *C. difficile* was isolated, all other specimens collected from that patient and their surroundings were also cultured on Cycloserine-Cefoxitin Fructose Agar with Horse Blood and Taurocholate (CCFA-HT) agar (Anaerobe Systems). Colonies resembling *C. difficile* (large, spreading, grey, ground glass appearance) were picked by a trained microbiologist and sub-cultured onto a blood agar plate (BAP). Growth from the subculture plate was identified using Matrix-assisted laser desorption/ionization-time of flight mass spectrometry (MALDI-TOF MS) (bioMerieux, Durham, NC). Upon identification, sweeps of *C. difficile* BAPs were collected in tryptic soy broth (TSB) and stored at -80C for sequencing. If both rectal swab sample and stool sample produced a *C. difficile* isolate, the stool isolate was preferentially used for analysis over the rectal swab isolate. The discharge / last specimen collected for an admission was also cultured for *C. difficile* if *C. difficile* was not isolated from the admission specimen. If *C. difficile* was isolated from the discharge/last specimen collected, then all specimens from that admission were also cultured for *C. difficile*.

*C. difficile* toxin enzyme immunoassay (EIA) was conducted as part of routine clinical care based on clinical suspicion of CDI. To be diagnosed with *C. difficile* infection (CDI), a patient must have been EIA+ for *C. difficile* toxin (Alere TOX A/B II); those who weren’t tested (due to no clinically significant diarrhea) or tested EIA- and were culture-positive for *C. difficile* were considered *C. difficile* carriers. Episodes of carriage or CDI are defined as the time from the first culture-positive specimen from a patient to the last culture-positive specimen during a given hospital admission.

### Short read sequencing and *de novo* genome assembly

Parameters used for computational tools are provided parenthetically. Total genomic DNA from *C. difficile* isolates was extracted from frozen plate scrapes using the QIAamp BiOstic Bacteremia DNA Kit (Qiagen) and quantified DNA with the PicoGreen dsDNA assay (Thermo Fisher Scientific). DNA from each isolate was diluted to a concentration of 0.5 ng/μL for library preparation using a modified Nextera kit (Illumina) protocol^28^. Sequencing libraries were pooled and sequenced on the NovaSeq 6000 platform (Illumina) to obtain 2D×D150Dbp reads. Raw reads were demultiplexed by index pair and adapter sequencing trimmed and quality filtered using Trimmomatic (v0.38, SLIDINGWINDOW:4:20, LEADING:10, TRAILING:10, MINLEN:60)^29^. Cleaned reads were assembled into draft genomes using Unicycler (v0.4.7)^30^. Draft genome quality was assessed using Quast^31^, BBMap^32^, and CheckM^33^, and genomes were accepted if they met the following quality standards: completeness greater than 90%, contamination less than 5%, N50 greater than 10,000 bp, and less than 500 contigs >1000bp.

### Isolate characterization and typing

A Mash Screen was used to identify likely related genomes from all NCBI reference genomes^34^. Average nucleotide identity (ANI) between the top three hits and the draft assembly was calculated using dnadiff^35^. Species were determined if an isolate had >75% alignment and >96% ANI^36^ to a type strain, and were otherwise classified as genomospecies of the genus level taxonomy call.

In silico multilocus sequence typing (MLST) was determined for all *C. difficile* and genomospecies isolates using mlst^37,38^. Isolate contigs were annotated using Prokka^39^ (v1.14.5, -mincontiglen 500, -force, -rnammer, -proteins GCF_000210435.1_ASM21043v1_protein.faa^40^). *cdtAB* was determined to be a pseudogene if there were three hits to *cdtB*, indicating the damaged structure of the pseudogene^41^. *C. difficile* clade was determined using predefined clade-MLST relationships described in Knight, et al^16^.

### Phylogenetic analyses

The .gff files output by Prokka^39^ were used as input for Panaroo (v1.2.10)^42^ to construct a core genome alignment. The Panaroo alignment was used as input to construct a maximum-likelihood phylogenetic tree using Fasttree^43^. The output .newick file was visualized using the ggtree (v3.4.0)^44^ package in R. Cryptic clade isolates were determined as such based on phylogenetic clustering with cryptic clade reference isolates.

### SNP analyses and network formation

We identified pairwise SNP distances between isolates identified as the same MLST type. The isolate assembly with the fewest number of contigs in an MLST group was chosen as a reference for that MLST group. Cleaned reads were aligned to their respective reference and SNP distances were calculated with snippy^45^. Pairwise SNP distances between isolates were calculated by merging VCF files with bcftools^46^ and a custom script. Only SNPs within the core genome of each MLST group were considered, thus core MLST SNPs were used for strain network determination. A cutoff of <=2 core MLST SNPs was used to define strain networks, as has been used previously to account for strain variation^15,47^.

### Phage identification and clustering

Isolate genomes were analyzed with Cenote-Taker 2^48^ to identify contigs with end features as direct terminal repeats (DTRs) indicating circularity, and inverted linear repeats (ITRs) or no features for linear sequences. Identified contigs were filtered by length and completeness to remove false positives. Length limits were 1,000 nucleotides (nt) for the detection of circularity, 4,000 nt for ITRs, and 5,000 nt for other linear sequences. The completeness was computed as a ratio between the length of our phage sequence and the length of matched reference genomes by CheckV^49^ and the threshold was set to 10.0%. Phage contigs passing these two filters were then run through VIBRANT^50^ with the “virome” flag to further remove obvious non-viral sequences^50^. Based on MIUViG recommended parameters^51^, phages were grouped into “populations” if they shared ≥95% nucleotide identity across ≥85% of the genome using BLASTN and CheckV.

### Analysis of genotypic associations with disease severity

Two previously sequenced retrospective cohorts from the same hospital were included to increase power^26,52^. In the analyses of toxigenic vs. nontoxigenic isolates from clade 1, pyseer^53^ was run using a SNP distance matrix (using snp-dist as above), binary genotypes (presence or absence of *tcdB*), and Panaroo-derived gene presence/absence data. In the analysis of CDI suspicion, all isolates from clade 1 were used that represented one isolate per patient-episode. Isolates recovered from environmental surfaces were excluded. Using these assemblies, a core genome alignment was generated using Prokka^39^ and Panaroo^42^ as above. SNP distances were inferred from the core-gene alignment using snp-dists^54^. Binary phenotypes were coded for the variable CDI suspicion, whereby isolates associated with a clinically tested stool were associated with symptomatic colonization (TRUE). Isolates that were associated with a surveillance stool and had no clinical testing associated with that patient timepoint were coded as non-symptomatic colonization (FALSE). Gene candidates filtered based on ‘high-bse’, and were annotated HMMER on RefSeq databases and using a bacteriophage-specific tool VIBRANT^50^. Selected outputs were visualized in R using the beta coefficient as the x-axis and the -log_10_(likelihood ratio test p-value) as the y-axis.

### Reference assembly collection

We chose 23 reference assemblies from Knight, et al^16^ for Figure 2c because of their MLST-clade associations (Supplementary Table 2). References span Clades 1-5 and cryptic clades C-1, C-2, and C-3, with one reference from each of the three most frequent MLSTs in each clade. Cryptic clade C-3 only had 2 reference assemblies available. References were annotated and included in phylogenetic tree construction as above.

All *Clostridioides difficile* genomes available on the National Institutes of Health (NIH) National Library of Medicine (NLM) were acquired for Figure 4c construction. References from NCBI (Supplementary Table 4) were included if they had less than 200 contigs. Assemblies that met these quality requirements were annotated and phylogenetically clustered as above.

## Results

### Surveillance of *C. difficile* reservoirs in hospital wards reveals patient colonization and environmental contamination

We prospectively collected patient and environmental samples to investigate genomic determinants of *C. difficile* carriage, transmission, and CDI (Figure 1). Across the study period, we enrolled 384 patients from 647 unique hospital admissions, and collected patient specimens upon admission and weekly thereafter (Supplementary Figure 1). We collected at least one specimen (clinical stool collected as part of routine care, study collected stool, or study collected rectal swab) from 364 admissions for a total of 1290 patient specimens (Table 1). We selectively cultured *C. difficile* from 151 stool specimens or rectal swabs if stool was unavailable or culture-negative. We also collected weekly swabs from the bedrails, sink surfaces, and in-room keyboards, for a total of 3045 swabs from each site. We cultured all environmental swabs collected from rooms in which patients that ever produced culturable *C. difficile* were housed, for a total of 398 swab sets plus one and two additional keyboard and sink handle swabs, respectively. In total, 22/398 (5.5%) of bedrail swabs cultured and 4/399 (1.0%) of keyboard swabs cultured were culture-positive for *C. difficile* (Figure 2a). *C. difficile* was never recovered from sink surfaces (all sinks on these units are hands-less activated) or other sampled sites. Collapsing multiple positive samples from the same patient admission results in 20 positive bedrails (20/79, 25.3% of all admissions with positive patient specimens) and 4 positive keyboards (4/79, 5.06% of all admissions with positive patient specimens) (Figure 2b).

**Figure 1:**
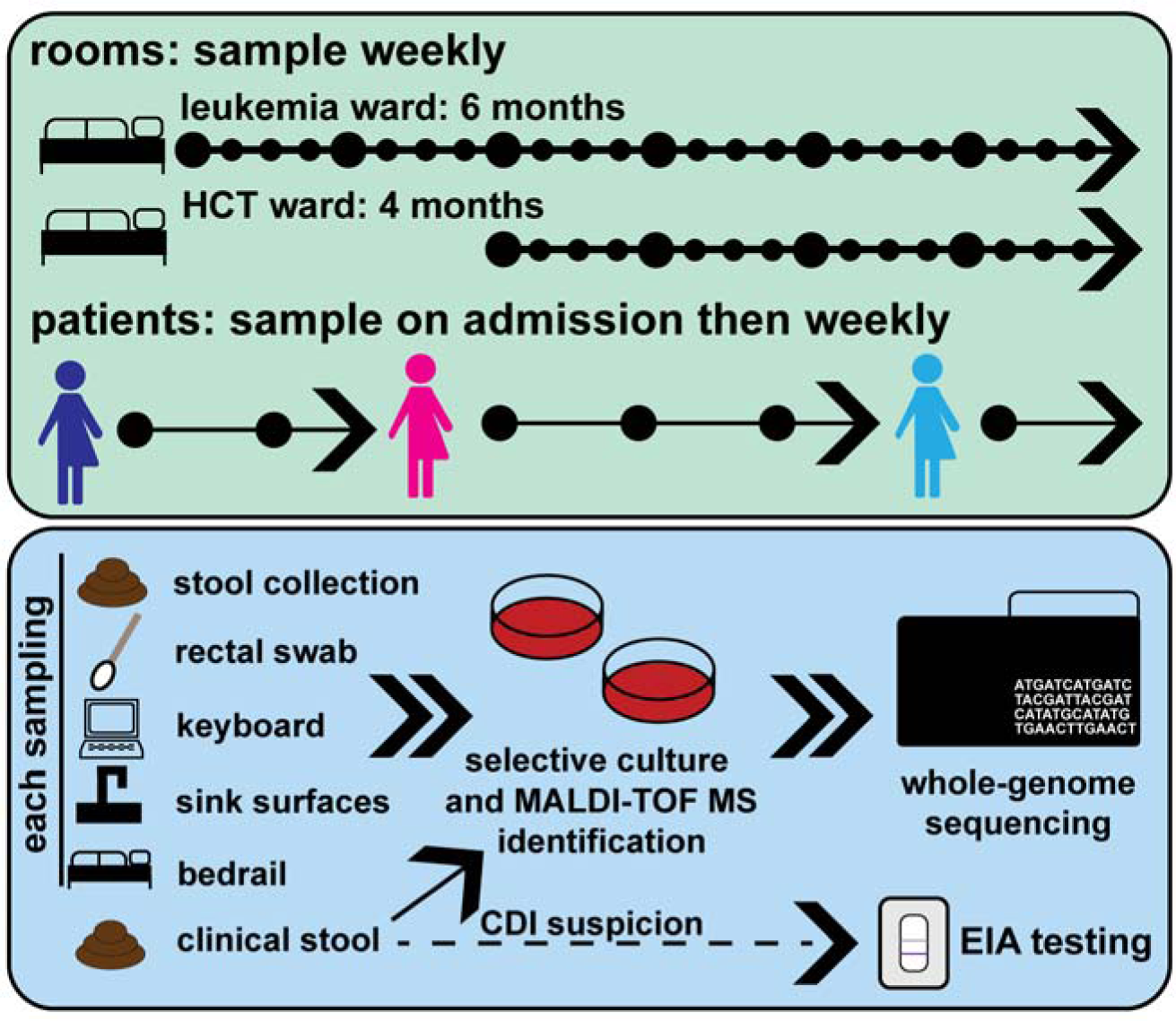
Study sampling and testing overview. Caption: a) We sampled a leukemia and hematopoietic stem cell transplant ward at Barnes-Jewish Hospital in St. Louis, USA for 6 and 4 months respectively. Patients were enrolled and sampled upon admission, and then weekly for their time in the study wards. Surfaces were sampled weekly across the duration of the study. All samples and stool collected as part of routine clinical care were subjected to selective culture and MALDI-TOF MS identification, and isolates were whole-genome sequenced. Results of EIA testing as part of routine care were obtained.

**Table 1.**
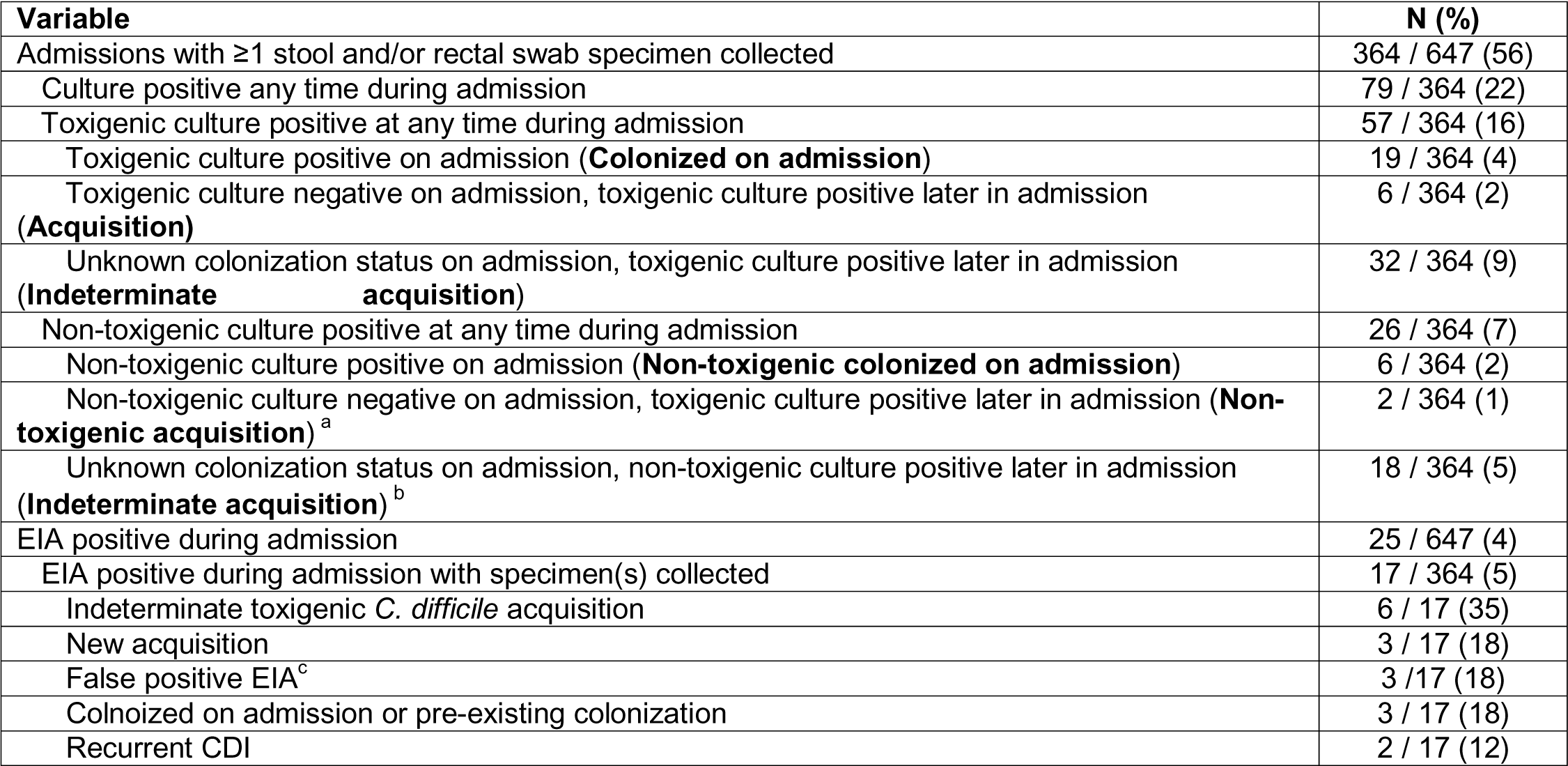
*C. difficile* epidemiology on the admission level (N=647 admissions)

Results from selective culture indicated that 21.7% of unique admissions (79/364 admissions with available specimens) were culture-positive for *C. difficile* at some point during their admission (Figure 2b, Table 1). Of these, 57 were toxigenic culture positive. 19 (4% of all admissions) patient-admissions were considered “colonized on admission” (i.e., toxigenic culture positive within the first three calendar days of admission), and toxigenic *C. difficile* was acquired in 6 (2%) admissions. For most toxigenic culture positive admissions (32; 9% of all admissions), *C. difficile* acquisition was considered indeterminate, meaning the earliest toxigenic culture positive specimen was collected on calendar day 4 or later during admission. Full admission-level culture results can be found in Table 1.

**Figure 2:**
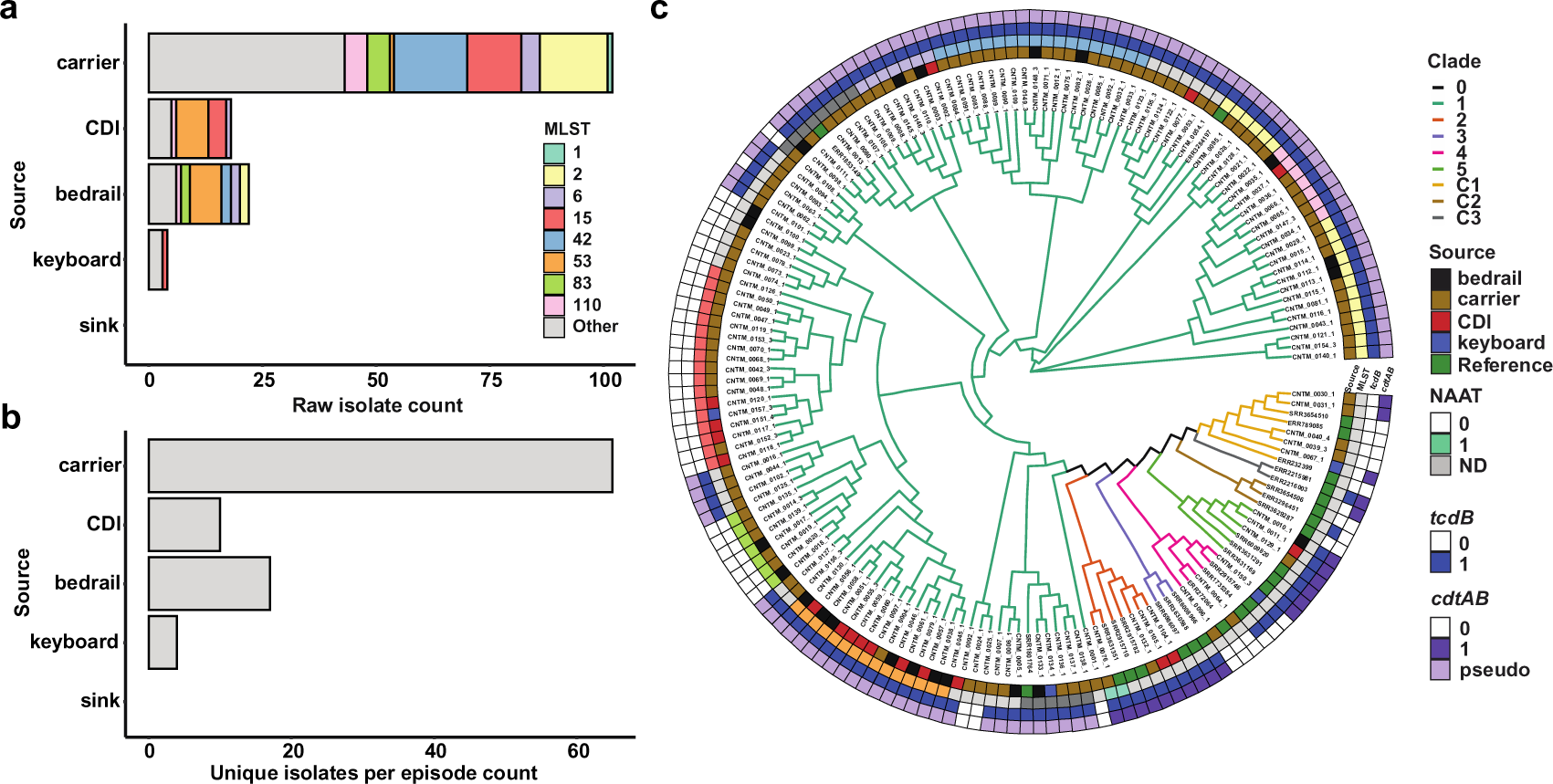
Total samples collected and phylogenetic relationships reveal carriers outnumber CDI patients and bedrails are the most commonly contaminated surface. Caption: Total a) isolates collected and b) culture-positive episodes from each source. We found more carriers than CDI patients, and bedrails yielded the most *C. difficile* isolates. c) Cladogram of all isolates collected during this study plus references.

### *C. difficile* carriers outnumbered patients with CDI

Patients with CDI were identified through routine clinical care, with CDI defined as patients who had stool submitted for *C. difficile* testing, as ordered by the clinical team when suspicious for CDI, and who tested positive for *C. difficile* toxins by enzyme immunoassay (EIA+). Otherwise, if they were culture positive and EIA- or culture positive and not EIA tested, they were considered carriers. Overall, 25 positive EIAs occurred during the study period; of these, 17 occurred during admissions with study specimens available for culture. Among these 17 admissions, 3 (18%) were considered new *C. difficile* acquisition; 6 (35%) had indeterminate timing of *C. difficile* acquisition; 3 (18%) were false positive EIAs, 3 (18%) were colonized on admission/pre-existing colonization, and 2 (12%) were recurrent CDI (Table 1). The substantial detection of longitudinal patient *C. difficile* colonization prompted us to investigate the genomic correlates of *C. difficile*-associated disease and transmission in these two patient populations.

### Phylogenetic clustering reveals lack of hypervirulent strains, presence of cryptic clades

We conducted whole-genome sequencing to ascertain phylogenetic distances among isolates and to identify closely related strains of *C. difficile.* We identified 141 isolate genomes as *C. difficile* (using a 75% alignment and 96% average nucleotide identity [ANI] threshold). One isolate was identified as *Clostridium innocuum* and five isolates were classified as *C. difficile* genomospecies (92-93% ANI). To contextualize population structure, we applied a previously established MLST-derived clade definition to our isolate cohort^16^. The majority of *C. difficile* isolates were from Clade 1 (131/146, 89.7% of *C. difficile* and genomospecies, Figure 2c). Four patient-derived isolates were identified from clade 2, but only one was of the hypervirulent strain ST1 (PCR ribotype 027)^6^. We found that the distribution of STs associated with carriers was significantly different from that of STs associated with CDI patients (p<0.001, Fisher’s exact test, Figure 2b) suggesting some strain-specificity to disease outcome.

Interestingly, the five genomospecies isolates clustered with other isolates belonging to a recently discovered *C. difficile* cryptic clade C-1 (Supplementary Figure 2). While cryptic clades are genomically divergent from *C. difficile*, these isolates can produce homologs to TcdA/B and cause CDI-like disease in humans^16,55^. In a clinical setting, they are frequently identified by MALDI-TOF MS as *C. difficile* and diagnosed as causative of CDI^55^. These data highlight the novel distribution of circulating *C. difficile* strains in the two study wards. While many patients with multiple isolates had homogeneous signatures of colonization (with closely related isolates), four patients (4/72 patients with positive cultures, 6%) produced isolates from distinct ST types.

### Carriers and CDI patients contribute to transmission networks and environmental contamination

Given the predominance of clade 1 isolates, we sought to identify clonal populations of *C. difficile* strains, indicative of direct *C. difficile* contamination (patient-environment) or transmission (patient-patient). We compared pairwise, core genome SNP distances within MLST groups to identify networks of transmission connecting isolates <=2 SNPs apart (Supplementary Figure 3). We identified a total of 25 strain networks, 2 of which contain patient isolates from different patients (networks 17 and 31, Figure 3a,d). These strain networks were temporally linked, as there were significantly fewer days between same-network isolates than isolates from different networks (p<2.2e-16, Wilcoxon, Figure 3b). We also sought to understand if CDI patients were more likely to contaminate bedrails than carriers. While we found slightly higher numbers of total bedrail isolates collected and unique bedrails contaminated by networks with CDI patients, neither comparison reached statistical significance (ns, Student’s t-test, Supplementary Figure 4a, b).

**Figure 3:**
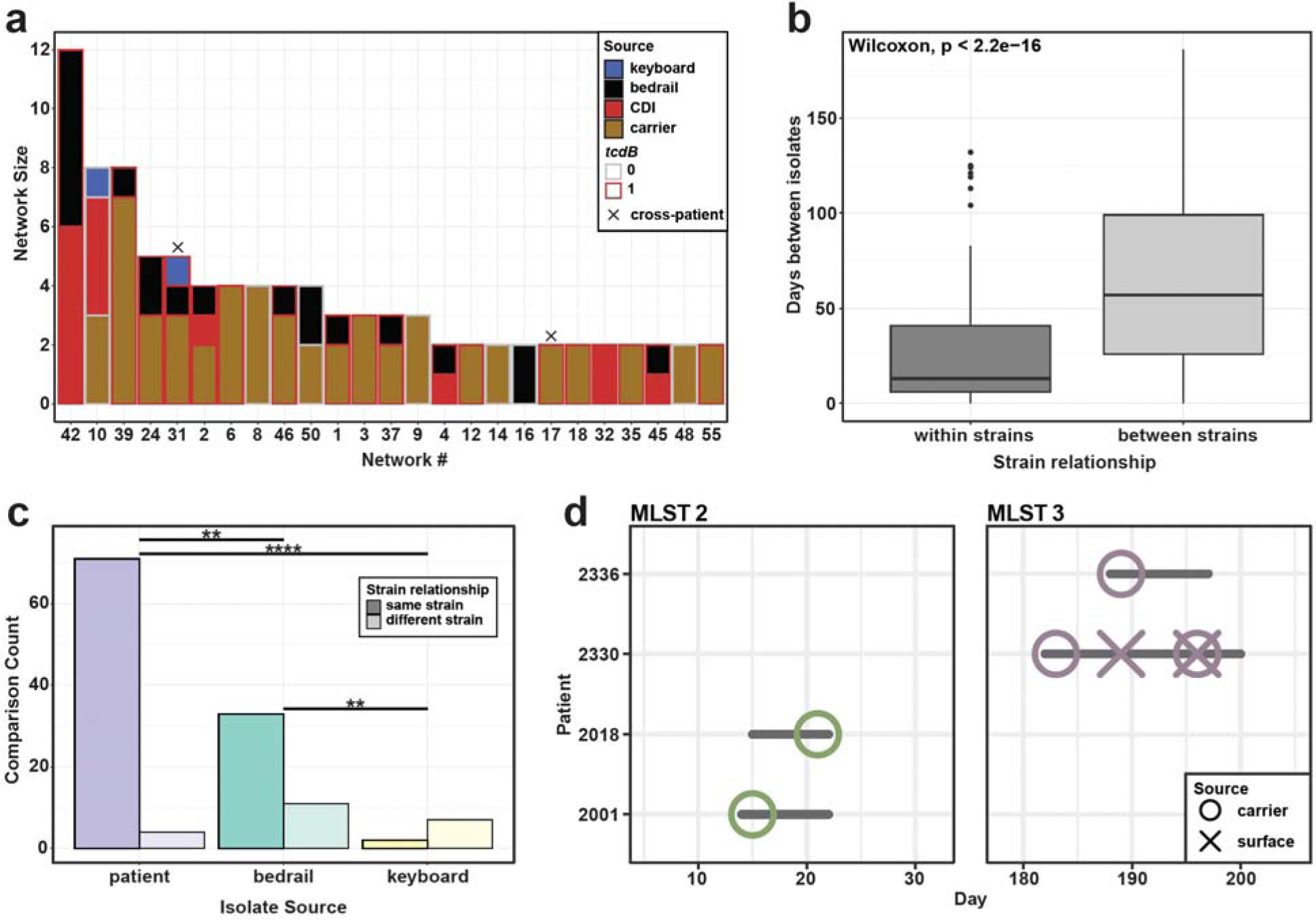
Surfaces are a site of environmental contamination and potential for transmission from colonized and CDI patients. Caption: a) Strain networks were defined by <=2 MLST core gene SNP cutoff. Network 10 includes the non-toxigenic isolates from Patient 2245 that are likely not responsible for the CDI. b) Absolute value of days between isolates within strains and between strains. Isolates within the same strain were significantly temporally linked (p<2.2e-16, Wilcoxon test). c) Number of comparisons in each group that fall within strain cutoff. Patient: between two isolates collected from the same patient; bedrail: between a patient isolate and an isolate taken from their bedrail; keyboard: between a patient isolate and an isolate taken from their keyboard. Fisher’s exact test, BH corrected. d) Strain tracking diagram of transmission networks associated with more than one patient. Colors indicate MLST of network and horizontal lines indicate stay in a room. Patient 2330 sheds *C. difficile* onto the bedrail and patient 2336 later is identified as a carrier of the same strain.

We compared strain connections among a single patient’s isolates from stool or rectal swab (‘patient’), and between these isolates and environmental isolates from their immediate surroundings (‘bedrail’ or ‘keyboard’, Figure 3c). While the majority of bedrail isolates fell within the same network as patient isolates from that room (33/44 comparisons, 75%), 25% (11/44 comparisons) were genomically distinct, suggesting contamination from alternate sources. Keyboards were mostly colonized with distinct strains from the patient (22%, 2/9 comparisons), indicating other routes of contamination (p<0.05, Fisher’s exact test, BH corrected. Figure 3c). Among the networks that contain multiple patients, we found no instances of potential transmission from the inhabitant of one room to the subsequent inhabitant. However, in both instances, each potential transmission is associated with a temporal overlap in patient stay in the same ward, providing epidemiological support for putative transmission (Figure 3d). Importantly, we found no networks connecting patients with CDI to *C. difficile* carriers, suggesting successful containment through contact precaution protocols. Two patients (Patients 2026 and 2056) carried a strain of *C. difficile* and later developed CDI with that same strain. These data suggest that direct transmission from CDI patients may no longer be the driving force behind patient CDI in this setting on contact precautions, and prompted us to investigate the relationship between isolate genetic diversity and patient symptomology.

### Accessory genomic elements are associated with host CDI symptoms

Despite evidence of transmission in this prospective study, a minority of patients were diagnosed with CDI relative to those asymptomatically colonized with *C. difficile* in part due to the presence of nontoxigenic *C. difficile* isolates (Figure 2b). To power our investigation of virulence determinants across patient-colonizing *C. difficile* strains, we performed whole genome sequencing on 102 additional patient-derived *C. difficile* isolates from a previously described *C. difficile*-colonized/CDI cohort from the same hospital^26^, where all patients had clinical suspicion of CDI (CDI suspicion), defined by a clinician ordering an EIA test during patient admission. Using an MLST-based clade definition as above, we identified that most CDI cases result from isolates within clade 1, though clade 2 isolates were more likely to be associated with CDI status (Figure 4a). The latter finding supports previous data indicating that clade 2 isolates are hypervirulent, often attributed to the presence of the binary toxin operon or increased expression from the PaLoc^19,56,57^. Meanwhile, some clade 1 isolates contain no toxin genes, indicating a diversity of colonization strategies in this lineage. Pangenomic comparison of nontoxigenic versus toxigenic isolates revealed that in addition to the PaLoc, the majority of our toxigenic isolates from clade 1 (95/131 of our cohort) possess remnants of the binary toxin operon (Figure 4b, *cdtR and cdtA/B* pseudogenes). Interestingly, we found that nontoxigenic isolates had a higher diversity of phage populations relative to toxigenic isolates (Supplementary Figure 5, p=5.7e-8, Wilcoxon). Given the previous report that full-length *cdtAB* was identified only within Clades 2, 3, and 5^16^, we investigated the conservation of *cdtR* (the transcriptional regulator of the binary toxin locus) across *C. difficile* strains (containing 5 lineages). We additionally examined >1400 *C. difficile* genome assemblies from NCBI (Supplementary Table 4, Figure 4c). *cdtR* (unlike *cdtAB*) was dispersed across clade 1 and significantly associated with *tcdB* (Figure 4d, Fisher’s exact test, BH corrected), suggesting a selective pressure to maintain some element of both toxin loci in these isolates. Notably, these operons are not syntenic, further underlining the significance of the association. From this association, we sought to further understand why some toxigenic clade 1 isolates cause CDI and some colonize without symptoms. Using 148 toxigenic clade 1 isolates collected from this study and two previous studies from the same hospital^26,52^, we utilized a bacterial GWAS approach, *pyseer*^53^, that identifies genetic traits associated with strains corresponding to patients with CDI symptoms. Using CDI suspicion (see Methods) as an outcome variable, we found that, multiple amidases (including *cwlD*), putative transcriptional regulators, and many genes of unknown function were enriched in isolates associated with CDI symptoms (Figure 4e). These data indicate that the most prevalent, circulating *Cd* strains that cause CDI are not the hypervirulent clade 2 strains, but highlight the possibility that remnant genomic features from epidemic strains and other features may contribute to virulence in this hospital clade of *C. difficile*.

**Figure 4:**
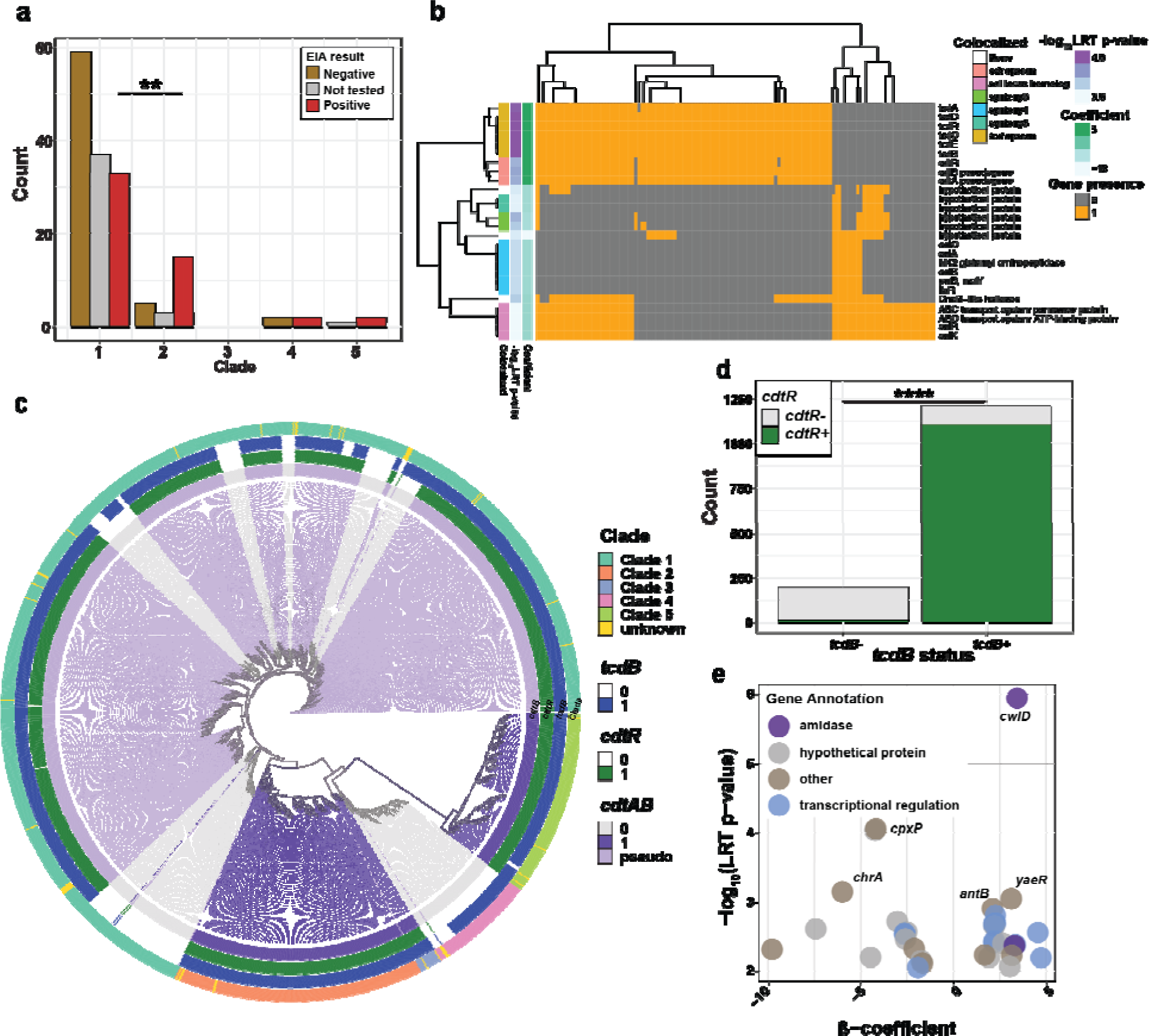
Clade 1 is responsible for the majority of CDI cases and carries unique correlates to symptom severity. Caption: a) EIA status by clade across this and a previous study^26^. Fisher’s exact test, p<0.01. b) Differentially abundant genes between toxigenic and nontoxigenic isolates in clade 1 from this study. Genes with a population structure adjusted p-value of <0.001 as produced by pyseer. c) Phylogenetic tree of >1400 *C. difficile* isolates from NCBI (Supplementary Table 4) depicting presence of binary toxin and PaLoc operons. d) Presence of full-length *cdtR* and association with *tcdB* presence. e) Filtered results (p-values <0.01) pyseer analysis evaluating gene association with CDI suspicion in Clade 1 isolates using the phylogenetically-corrected p-values (LRT). Purple color indicates p<0.001. Positive beta coefficient indicates gene association with CDI suspicion, while negative beta indicates asymptomatic colonization.

## Discussion

Through our prospective genomics study of two hospital wards, we were able to identify connections between contamination of different surfaces and the strains carried by hospitalized patients and quantify some spread between carriers. Our estimates of the prevalence of patients with CDI (3.8%) agree with other estimates of 2-4% CDI in patients with cancer^58–60^. While many studies have quantified surface contamination, few have had the genomic resolution to identify clonality between isolates indicating transmission or patient shedding^61–63^. We observed distinct contamination between a patient’s bedrail that differed from the strain the patient themselves carried, indicating that the bedrail may be a point of transmission. Further, we did not identify any instances of CDI that could be genomically linked to an earlier CDI case or *C. difficile* carrier. We identified two possible instances of transmission between carriers, though neither of these occurrences resulted in CDI. As this finding is in the context of contact precautions for CDI patients, it indicates that these strategies are successful at limiting transmission of *C. difficile* that causes CDI, and there is limited risk of CDI due to transmission from carriers. These findings confirm previous suggestions that carriers are not a significant risk for transmission leading to CDI^64,65^.

Our data suggests the need to investigate diverse lineages of *C. difficile* beyond previous epidemic strains to clarify mechanisms of disease. Among 79 culture positive admissions, we only isolated the epidemic PCR ribotype 027 strain once, causing just one case of CDI within our cohort. Because the overall burden of Clade 1 isolates was so high, we hypothesize that understanding the mechanisms and genomic factors by which these isolates cause disease may become more important as the burden of PCR ribotype 027 decreases^66^. While Clade 1 isolates associated with CDI symptoms are expectedly toxigenic (containing the toxin genes in the PaLoc), we also found an enrichment in two different amidase genes, that could either contribution to differences in germination rate or possess endolysin function^67,68^. How the function of such a gene contributes to an increase in symptomology remains to be understood. Further, we confirmed a genetic relationship between *cdtR* and *tcdB* across *C. difficile* lineages that indicates some evolutionary pressure for maintaining the regulatory gene of the less prevalent toxin operon (*cdtR*). This phylogenomic analysis supports recent functional data from clade 2 isolates that the presence of *cdtR* increases the expression of *tcdB* disease severity in an animal model of CDI^57^. While this was previously suggested *in vitro*, it is unclear how generalizable this relationship is across lineages^56^. In fact, we predict that clade 1 isolates containing only *cdtR* and the PaLoc may produce more toxin *in vivo* than those without *cdtR*. Future studies are warranted to investigate the role of both classes of genes implicated in this phenotype.

Our study has a number of important limitations. As this study focused on *C. difficile* colonization, disease, and transmission in two wards in the same hospital system, studies with increased sample size or meta-analysis studies are necessary to understand generalizable epidemiological measurements of *C. difficile*-patient dynamics^15^. For example, we were unable to fully quantify in-unit transmission, as not all patients were able to provide stool specimens and/or consented to rectal swabs within 3 days of admission. Additionally, since we did not culture all environmental swabs or specimens, we likely missed some instances of surface contamination or more transient patient carriage, and thus expect that we underestimated the frequencies of contamination and carriage in these wards. Further, the patients housed in the leukemia and HCT ICUs are unique due to their long hospital stays and high antibiotic exposure^69^. While this population was selected specifically to allow us to increase our sample sizes, these patient characteristics could contribute to extended *C. difficile* colonization time relative to other hospital patient cohorts. Finally, we note the evidence for multi-strain colonization within a single patient (Patient 2330). This patient was diagnosed with CDI, but only nontoxigenic *C. difficile* was isolated (network 10). This could be due to co-colonization, and we never isolated the toxigenic isolate responsible for the CDI, or a false positive toxin EIA. Given our approach of only culturing and sequencing single isolates per patient timepoint, future studies are needed to investigate the extent of within-patient *C. difficile* strain diversity by interrogating additional cultured isolates per samples or via metagenomic methods^70^.

Despite these limitations, this work highlights new investigative directions for the prevention of CDI. This work and others find risk for patients carrying *C. difficile* long-term in development of CDI, and we hypothesize that the mechanisms of virulence may be more complex than previous epidemic strains. We also hypothesize that non-CDI carriers contribute to the expansion of *C. difficile* transmission networks and emphasize the need to update infection prevention efforts as this landscape evolves. Indeed, though much human and animal research has focused on epidemic strains that are two decades old, we and others have identified more disease and colonization, largely from clade 1 lineages. We also investigate gene flux of phage like elements, that may play an important role in colonization, particularly in nontoxigenic isolates. Moreover, within this lineage we found a mosaic representation of genes associated with the PaLoc that highlight the possibility of different mechanisms of colonization and virulence by this population of *C. difficile*. Future studies utilizing other human cohorts or animal models are warranted to investigate disease and pathogenicity caused by Clade 1 *C. difficile* strains.

## Conclusions

Our study provides new insight into the nature of prevalent *C. difficile* strains in a hospital setting, transmission between carriers, and pathogen evolution during circulation. Longitudinal sampling of surfaces and patient stool revealed that both toxigenic and nontoxigenic strains of *C. difficile* clade 1 are prevalent in these two wards. Moreover, our estimation of carriage patterns emphasizes the need for further investigation into longitudinal carriage of *C. difficile* and its increased risk for CDI. We also note distinct differences in phage carriage between toxigenic and nontoxigenic *C. difficile*. We identity novel associations of accessory genes with CDI symptomology and toxigenicity (*cdtR* and *cwlD*). Our data highlights the complexities of understanding disease from this pathogen in a hospital setting and the need to investigate mechanisms of *in vivo* persistence and virulence of prevalent lineages in the host gut microbiome.

## Data Availability

The datasets generated and analyzed during the current study are available in NCBI GenBank under BioProject accession no. PRJNA980715.

## List of Abbreviations

BAP: blood agar plate
CCFA-HT: Cycloserine-Cefoxitin Fructose Agar with Horse Blood and Taurocholate
CCMB-TAL: Cycloserine Cefoxitin Mannitol Broth with Taurocholate and Lysozyme
CDI: *Clostridioides difficile* infection
EIA: enzyme immunoassay
HAI: healthcare-associated infection
HGT: horizontal gene transfer
MALDI-TOF MS: Matrix-assisted laser desorption/ionization-time of flight mass spectrometry
NTCD: non-toxigenic *C. difficile*
PaLoc: pathogenicity locus TCD, toxigenic *C. difficile*
TSB: tryptic soy broth

## Declarations

### Ethics approval and consent to participate

The study protocol was approved by the Washington University Human Research Protection Office (IRB #201810103). All participants provided written informed consent.

## Consent for publication

### Not applicable

#### Competing interests

The authors declare that they have no competing interests.

### Funding

This work was supported in part by an award to ERD and GD through the Foundation for Barnes-Jewish Hospital and Institute of Clinical and Translational Sciences. This publication was supported by the NIH/National Center for Advancing Translational Sciences (NCATS), grant UL1 TR002345 (PI: B. Evanoff). This work was also supported by funding through the CDC BAA #200-2018-02926 under PI Erik Dubberke. SRSF is supported by the National Institute of Child Health and Human Development (NICHD: https://www.nicdhd.nih/gov) of the NIH under award number T32 HD004010 (PI: P. Tarr). The conclusions from this study represent those of the authors and do not represent positions of the funding agencies.

### Authors’ contributions

SRSF, KAR, ERD, and GD participated in idea formulation and funding for this project. TH, KAR, CC, ZHI, ELS, and ERD conducted participant enrollment, sample collection, and microbiological isolation. EPN, SRSF, KZ, and GD conducted all sequencing analysis and figure generation. EPN and SRSF completed the writing of the manuscript. All authors read and approved the final manuscript.

## Acknowledgements

The authors are grateful for members of the Dantas lab for their helpful feedback on the data analysis and preparation of the manuscript. The authors would also like to thank the Edison Family Center for Genome Sciences and Systems Biology staff, Eric Martin, Brian Koebbe, MariaLynn Crosby, and Jessica Hoisington-López for their expertise and support in sequencing/data analysis.

## Supplemental Figures and Tables Titles and Captions

**Supplementary Figure 1:**
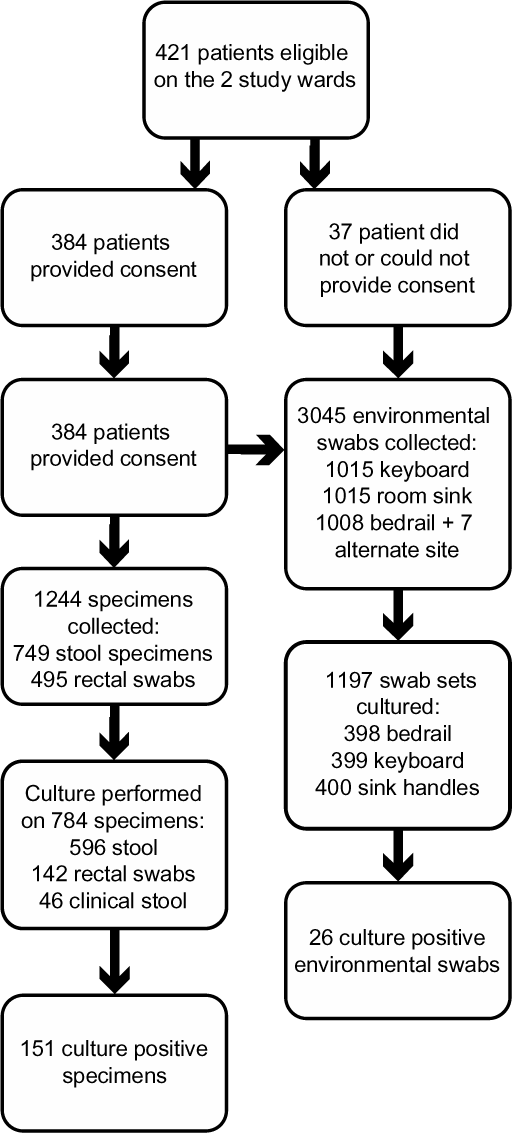
Bubble plot of enrollment, collection, and culture numbers.

**Supplementary Figure 2:**
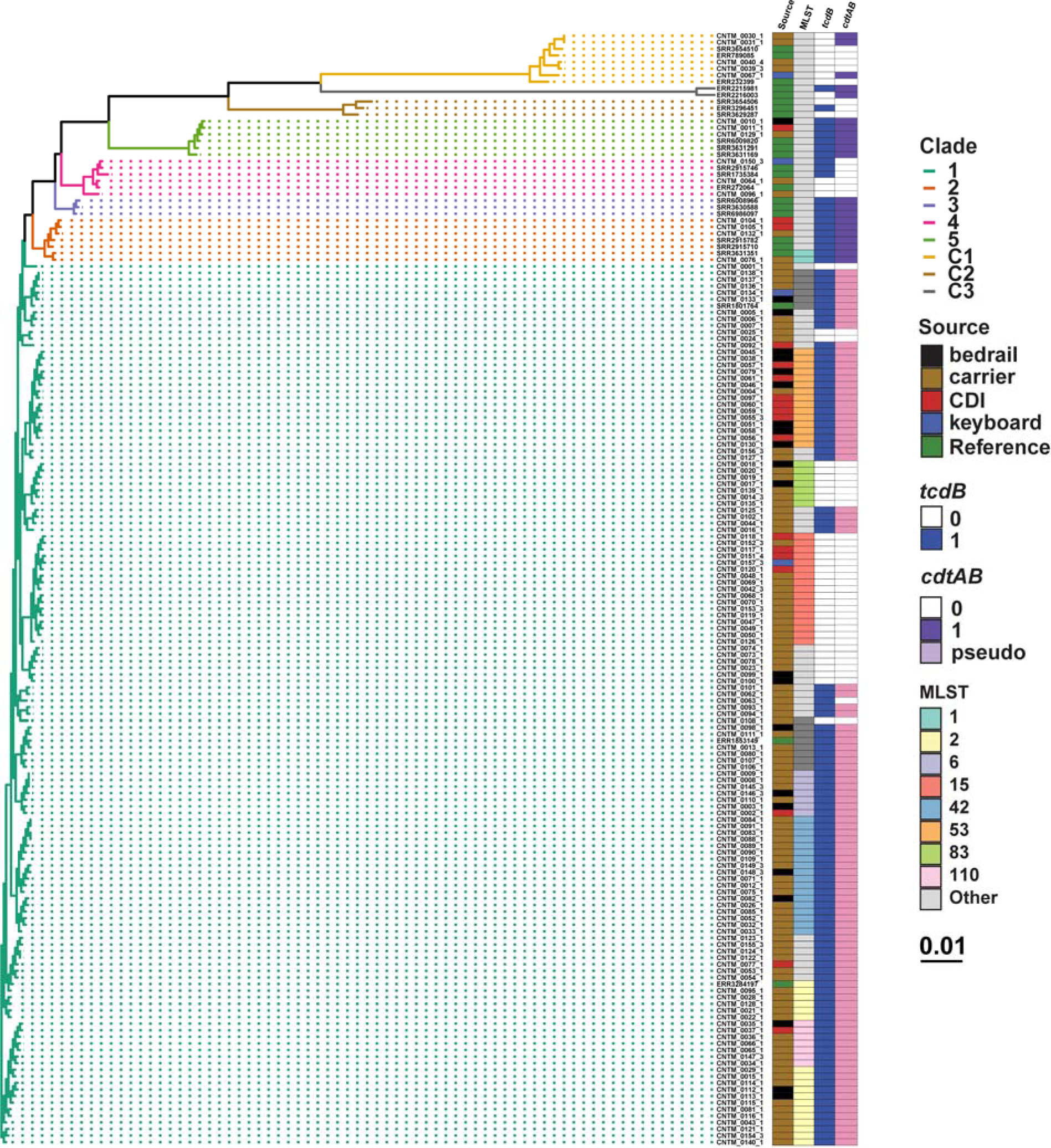
Phylogenetic tree of isolates collected in this study and select references (Supplementary Table 2).

**Supplementary Figure 3:**
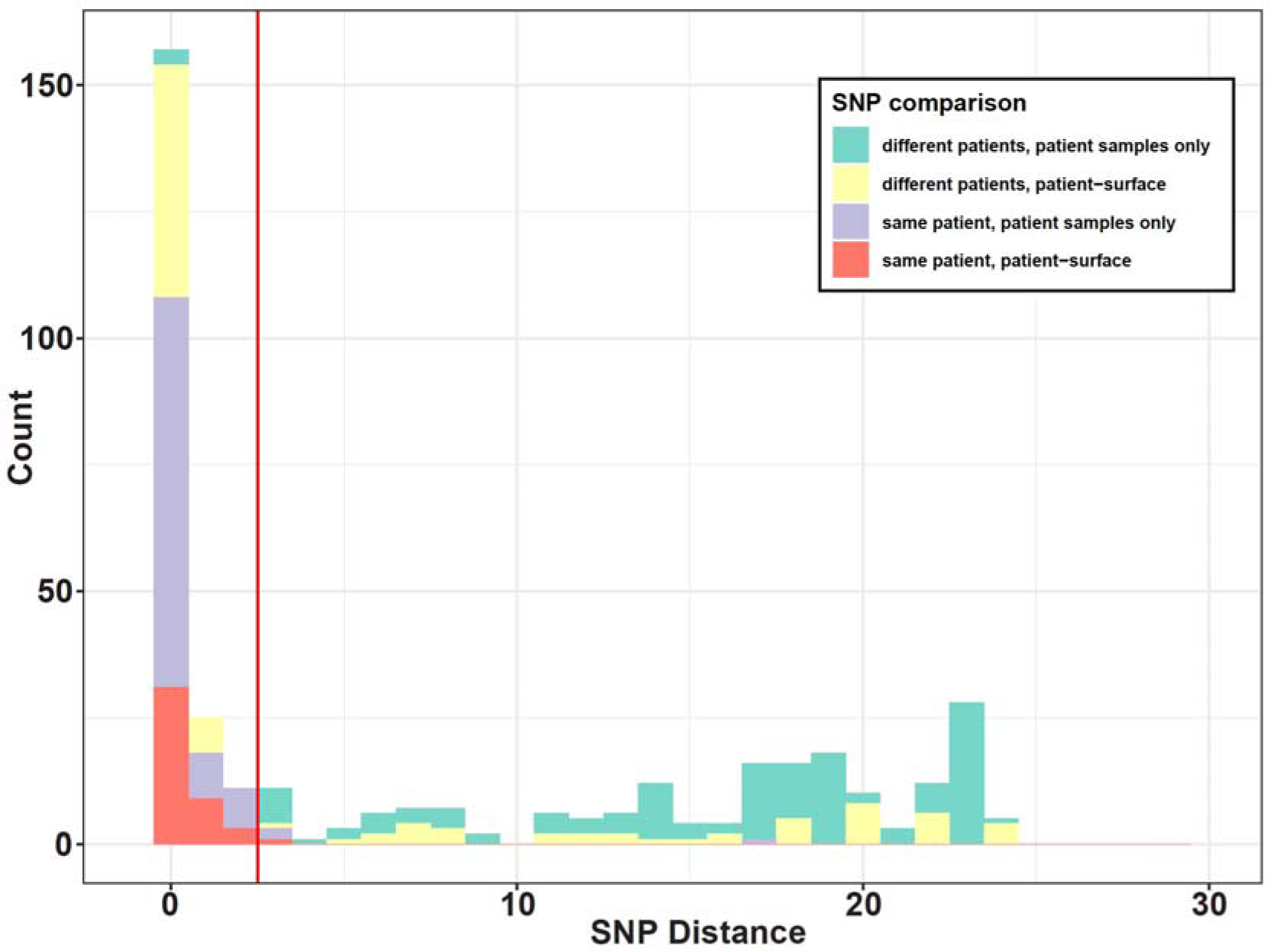
Histogram of core genome SNP distances between different within-MLST isolate comparisons, zoomed to show SNP cutoff (red line at 2 SNPs).

**Supplementary Figure 4:**
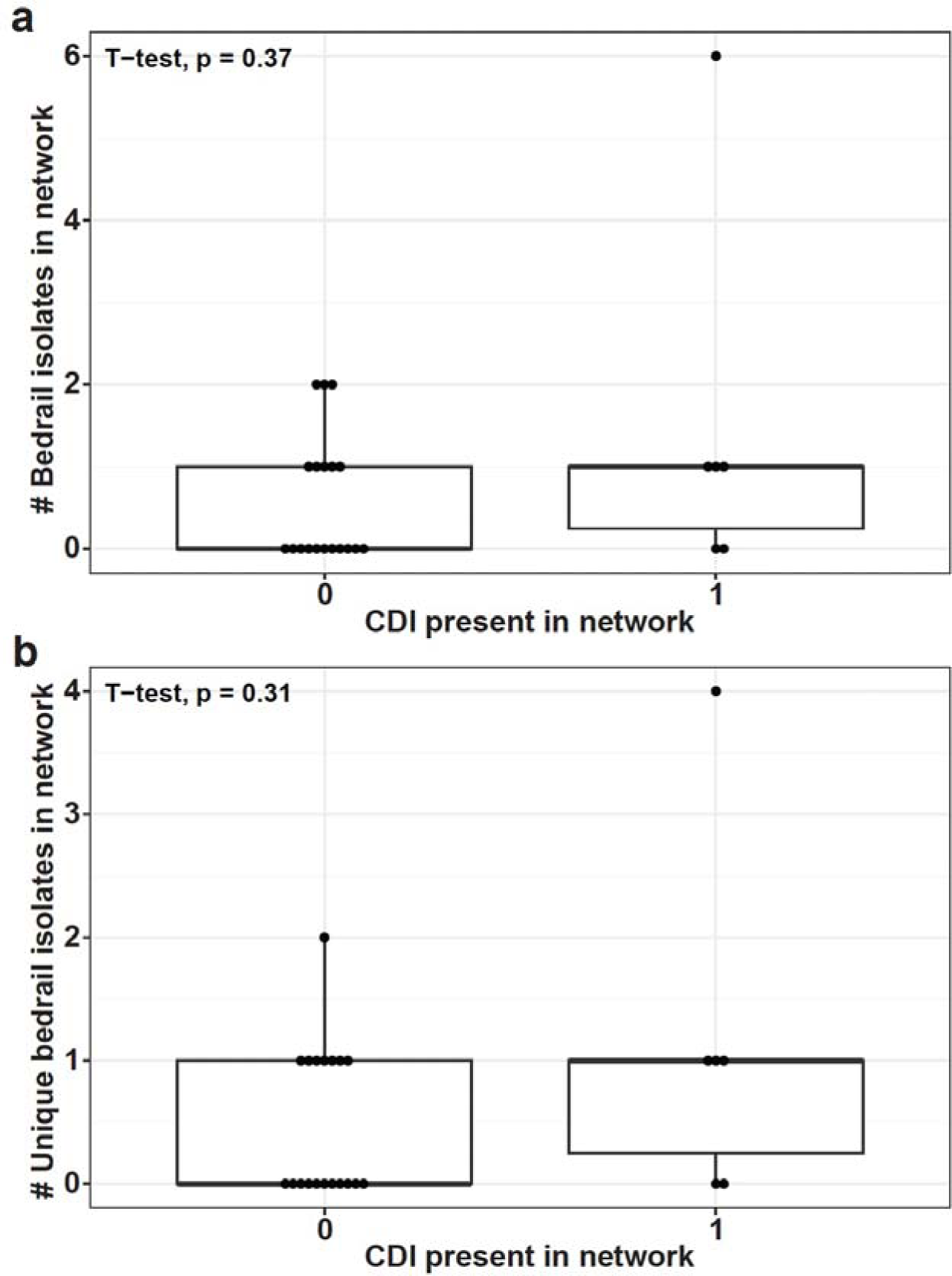
A) Total number of bedrail isolates in networks either containing a CDI case or not containing a CDI case. B) Number of unique bedrails contaminated in a network either containing a CDI case or not containing a CDI case. Student’s t-test.

**Supplementary Figure 5:**
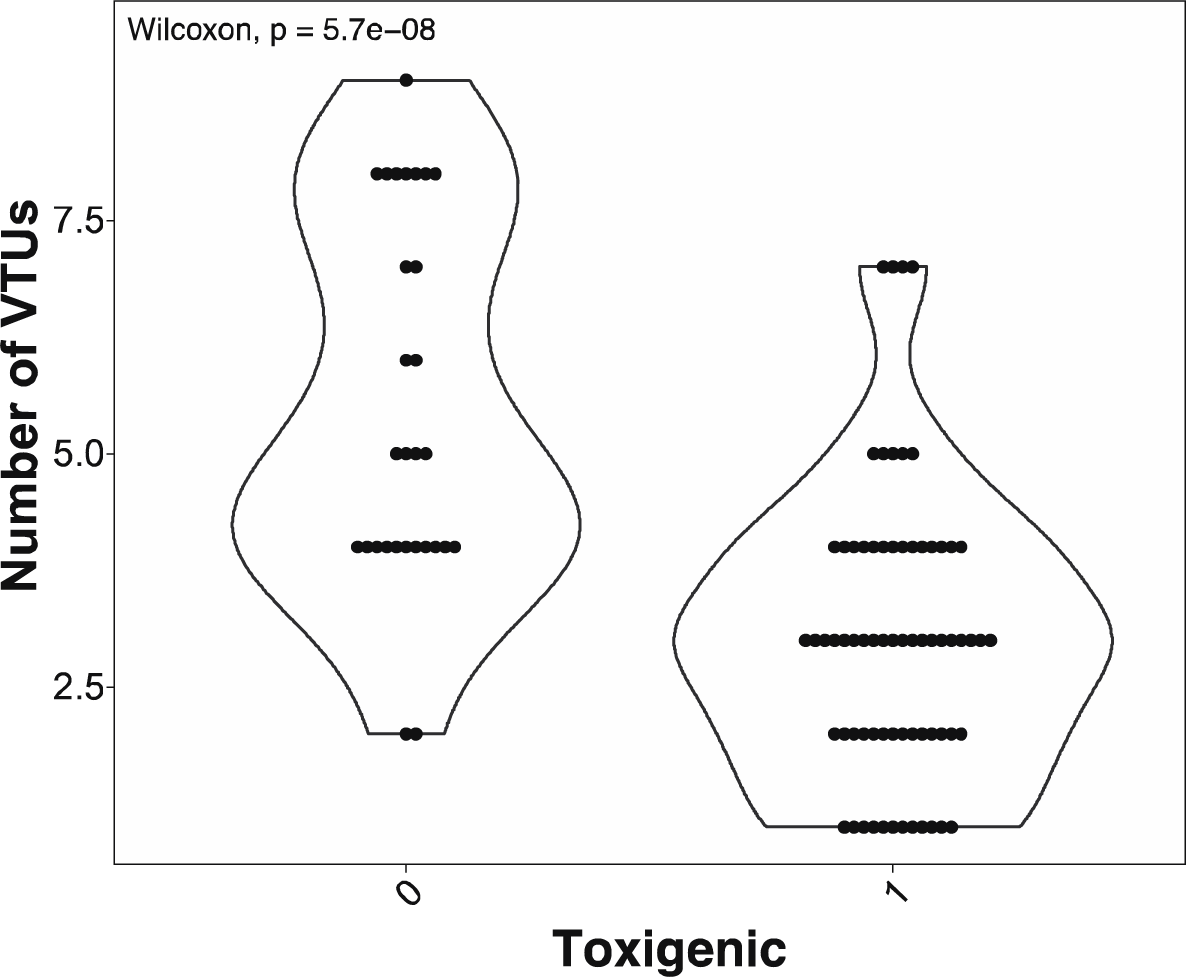
Phage population richness across toxigenic and nontoxigenic isolates in our cohort. Wilcoxon test, p=5.7e-8.

## Notes

### Competing Interest Statement

The authors have declared no competing interest.

### Summary of Updates

Figure 2, Figure 3, and Figure 4 revised

